# Growth in Childhood and Cardiovascular Health in Young Adulthood in Four Low- and Middle- Income Countries: Findings from the COHORTS Consortium

**DOI:** 10.64898/2026.08.13.26360417

**Authors:** Sarah Qabazard, Lisa Ware, Bernardo Lessa Horta, Natalia Peixoto Lima, Fernanda Kroker, Manuel Ramirez-Zea, Delia B. Carba, Isabelita Bas, Judith Borja, Linda Adair, Nanette Lee, Tita Lorna Perez, Linda M Richter, Shane Norris, David Flood, Darwin Labarthe, Aryeh D Stein

**Affiliations:** Nutrition and Health Sciences Program, Laney Graduate School, Emory University, Atlanta, GA, USA; South African Medical Research Council Ageing African Adult Research Unit, School of Clinical Medicine, Faculty of Health Sciences, University of the Witwatersrand, Johannesburg, South Africa; Postgraduate Program in Epidemiology, Universidade Federal de Pelotas, Brazil; INCAP Research Center for the Prevention of Chronic Diseases, Institute of Nutrition of Central America and Panama, Guatemala City, Guatemala; USC-Office of Population Studies Foundation, Inc., University of San Carlos, Cebu, Philippines; UNC Chapel Hill, Carolina Population Center, Chapel Hill, NC, USA; School of Human Development and Health, University of Southampton, UK; Department of Medicine, University of Michigan, USA; Department of Preventive Medicine, Feinberg School of Medicine, Northwestern University, Chicago, IL, USA; Hubert Department of Global Health, Rollins School of Public Health, Emory University, Atlanta, GA, USA

**Keywords:** Cardiovascular Health, Child Growth, Early-life Factors, Low- and Middle-Income Countries

## Abstract

**Background:** Early-life growth is associated with individual cardiometabolic risk factors, but its relationship with overall cardiovascular health (CVH) in low- and middle-income countries (LMICs) is unclear. We examined associations of maternal, household, and child growth factors with young-adult CVH across four LMIC birth cohorts.

**Methods:** We analyzed harmonized data from the Consortium of Health-Oriented Research in Transitioning Societies (COHORTS), including 4,582 participants ages 18-30 years from Brazil, Guatemala, the Philippines, and South Africa. CHV was assessed using a modified American Heart Association Life’s Simple 7 score based on body mass index (BMI), blood pressure (BP), fasting blood glucose (FBG), and smoking. Site-specific multivariable ordinal logistic regression models evaluated associations between early-life factors and CVH.

**Results:** Men had poorer CVH than women across most sites, largely because of less favorable BP and smoking profiles. Higher birthweight was associated with lower odds of better CVH in Brazil (AOR=0.81; 95% CI: 0.71-0.94) and the Philippines (AOR=0.63; 95% CI: 0.45-0.87). Greater conditional relative weight at 2 years was also inversely associated with CVH in both sites. Birthweight, conditional height and conditional relative weight at 2 years were strongly associated with adult BMI, whereas associations with BP and FBG were weaker. Attained schooling was associated with CVH in Brazil (AOR = 1.13 per year; 95% CI: 1.10–1.16), and the Philippines (AOR = 1.17; 95% CI: 1.10–1.24).

**Conclusions:** Early-life growth patterns and educational attainment are associated with cardiovascular health in young adulthood across diverse LMIC settings, supporting life-course strategies to promote cardiovascular health.

## INTRODUCTION

Cardiovascular disease (CVD) is the leading cause of death globally, accounting for 32% of all deaths, with nearly 80% occurring in low- and middle-income countries (LMICs) (1, 2, 3). This disproportionate burden underscores significant global health disparities, as LMICs often face unique challenges, including limited access to healthcare, under-resourced prevention programs, and the simultaneous burden of communicable and non-communicable diseases (4). Addressing these disparities requires a deeper understanding of cardiovascular health (CVH) in LMIC contexts.

Numerous studies have explored links between early-life factors, such as maternal and household socioeconomic status, child growth, and cognitive development, and later-life outcomes (5, 6). The first 1000 days of life, spanning from conception to a child’s second birthday, represent a critical window for growth and development, with long-term implications for health outcomes, including CVH (7). For instance, size at birth, weight gain, stature, and early-life poverty have been associated with school attainment, wealth, cognitive functioning, and specific cardiometabolic risk factors (7, 8, 9, 10, 11, 12, 13). However, much of this literature has focused on individual cardiometabolic risk factors rather than a holistic measure of CVH.

The American Heart Association’s Life’s Simple 7 (LS7) metric offers a comprehensive approach to assessing CVH by integrating seven key components: diet, physical activity, body mass index, blood pressure, blood glucose, total cholesterol, and smoking (14). Despite its potential to guide prevention strategies, LS7 has primarily been studied cross-sectionally in high- and low- and middle-income countries, limiting its ability to capture the long-term effects of early-life factors on CVH (15, 16).

The Consortium of Health-Oriented Research in Transitioning Societies (COHORTS) collaboration provides a unique opportunity to explore these relationships across distinct LMIC populations (17). COHORTS pools data from prospective birth cohorts from Brazil, Guatemala, India, Philippines, and South Africa, offering a rich longitudinal dataset on early-life exposures and their associations with young-adult health indicators (18, 19, 20, 21). This diversity allows for a comprehensive examination of how maternal and child and socioeconomic factors interact to influence CVH across life stages.

Previous analyses using COHORTS data have shown that early growth patterns, such as birthweight and weight-for-age at 2 years, are associated with individual adult cardiometabolic risk factors, including body mass index, blood pressure, and glycemia (9, 12). For instance, higher birthweight and conditional relative weight at 2 years were associated with increased adult fat mass and fat-free mass (9). However, it remains unclear how these findings relate to overall cardiovascular health. The aim of this study is to examine the associations of early-life exposures, including maternal and household factors and child growth indicators, with cardiovascular health in young adulthood. This study will help inform public health policies that promote healthy growth and education in LMICs, enhancing lifelong cardiovascular health and reducing the global CVD burden.

## METHODS

### Study design and participants

We conducted a secondary analysis of COHORTS data, a consortium of prospective birth cohorts from five LMICs. This study includes data from four of the cohorts: individuals born: 1) in 1982 Pelotas, Brazil (Pelotas Birth Cohort); 2) between 1962 and 1977 in four villages in Guatemala (Institute of Nutrition of Central America and Panama (INCAP) Nutrition Supplementation Trial Cohort); 3) between 1983 and 1984 in Cebu, the Philippines (Cebu Longitudinal Health and Nutrition Survey - CLHNS); and 4) in 1990 in Soweto or Johannesburg, South Africa (Birth to Twenty BT20). Detailed profiles of these cohorts (hereon referred to as sites) are described elsewhere (18, 19, 20, 21). While these cohorts provide valuable longitudinal data, they reflect specific populations within each country and are not nationally representative. All fieldwork adhered to procedures approved by local ethics review boards, and written informed consent was obtained from all participants or their parents, as applicable. This analysis was approved by the Emory University Ethical Review Board (Ref: 95960).

Figure 1 shows the flow chart of the analytic sample selection for the sites included in this analysis. We excluded participants who were pregnant at time of anthropometric assessment and those with missing early-life exposures, CVH indicators or covariates. Each site was analyzed independently.

**Figure 1.**
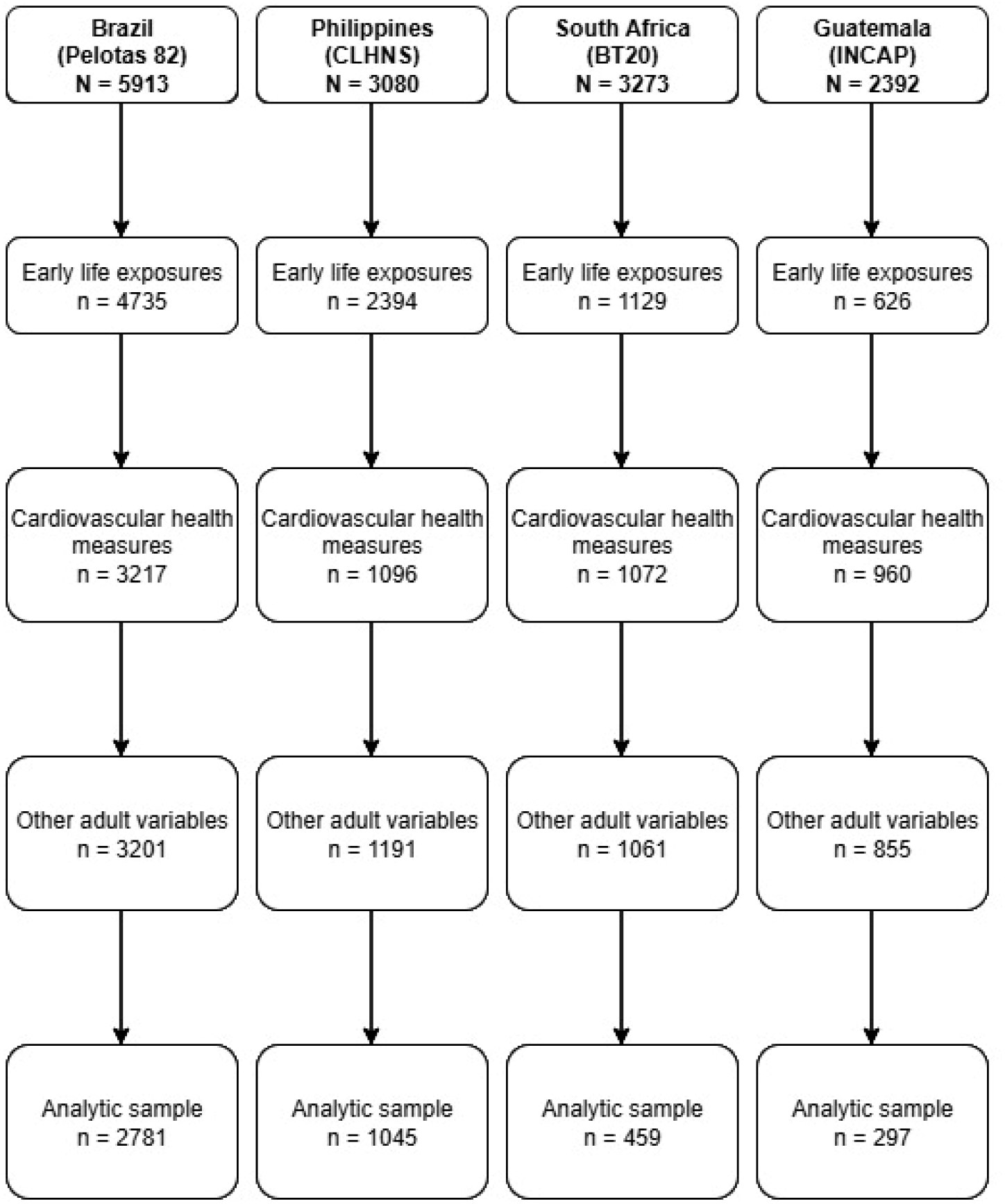
Flow chart of data completeness to reach the analytic sample for each Consortium of Health-Oriented Research in Transitioning Societies (COHORTS) site. Pelotas 82: 1982 Pelotas, Brazil Birth Cohort. CLHNS: Cebu Longitudinal Health and Nutrition Survey. BT20: Soweto/ Johannesburg, South Africa Birth to Twenty cohort. INCAP: Institute of Nutrition of Central America and Panama Nutritional Trial Cohort.

### Variable specifications

Early-life exposures were grouped into two sets to explore the association with young adulthood (defined as 18-30 years) CVH, as depicted in Figure 2. Set 1 (Maternal/ Household factors) includes maternal age (which influences pregnancy and developmental risks), maternal height (reflecting nutritional and health status), maternal schooling (indicative of health practices and resource access), birth order and household wealth (overall living conditions and healthcare access). Set 2 (Child size) includes birthweight, length-for-age Z score at 2 years and weight-for- age Z score at 2 years, all crucial predictors of future adult health. These exposures were selected for their potential link to CVH (14). Statistical methods were used to isolate the effects of linear growth and weight gain due to their strong correlation with each other. Specifically, we derived conditional height-for-age and conditional relative weight at age 2 years, through regression analysis, within each site (12). The residuals from these models were used in our analytic models.

**Figure 2.**
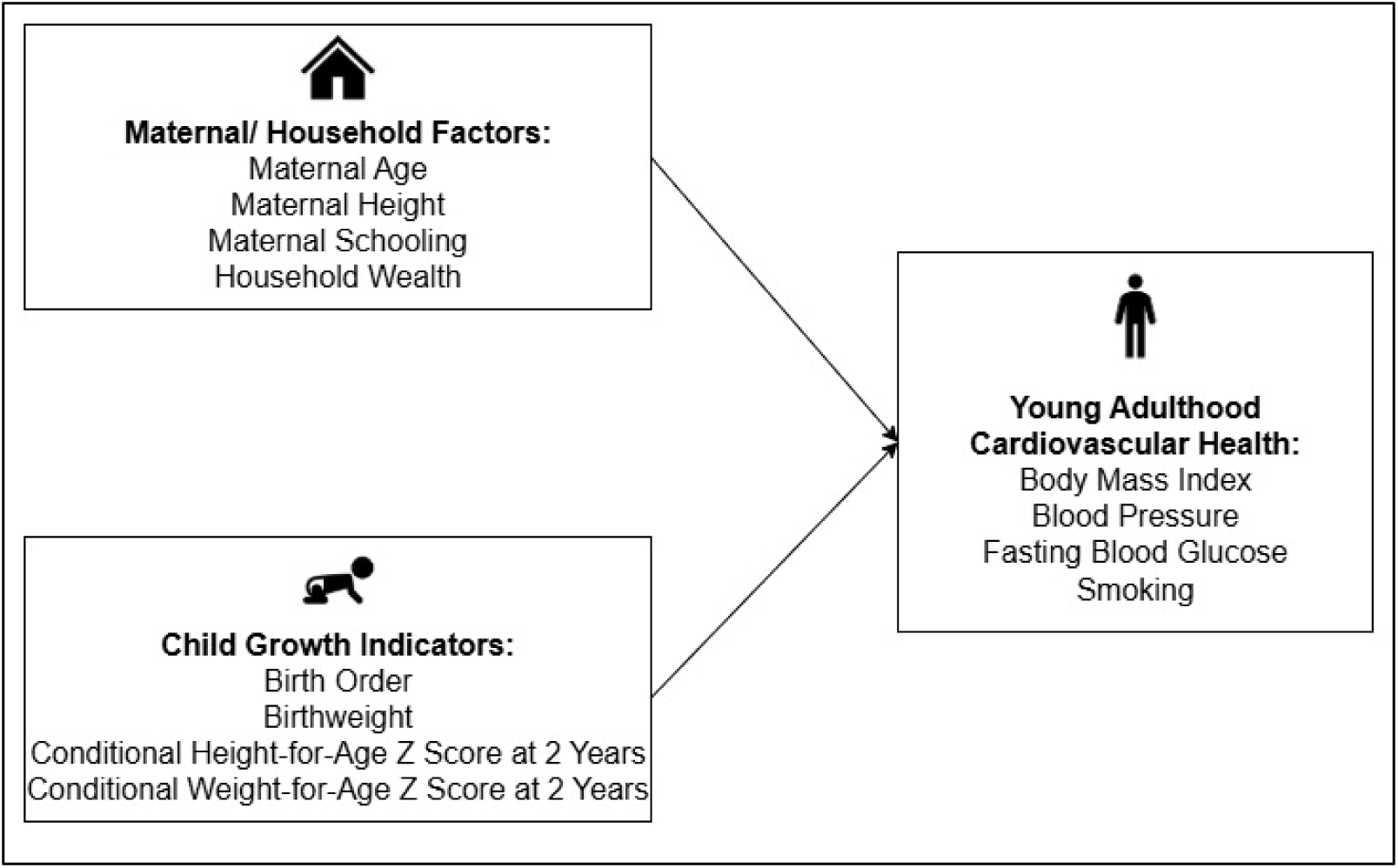
Conceptual framework of early-life predictors and cardiovascular health later in life. The framework highlights maternal and household factors and child growth indicators as key exposures. These factors may be associated with total cardiovascular health and/or key components, including: body mass index, blood pressure, fasting blood glucose, and smoking status in young adulthood.

**Figure 3.**
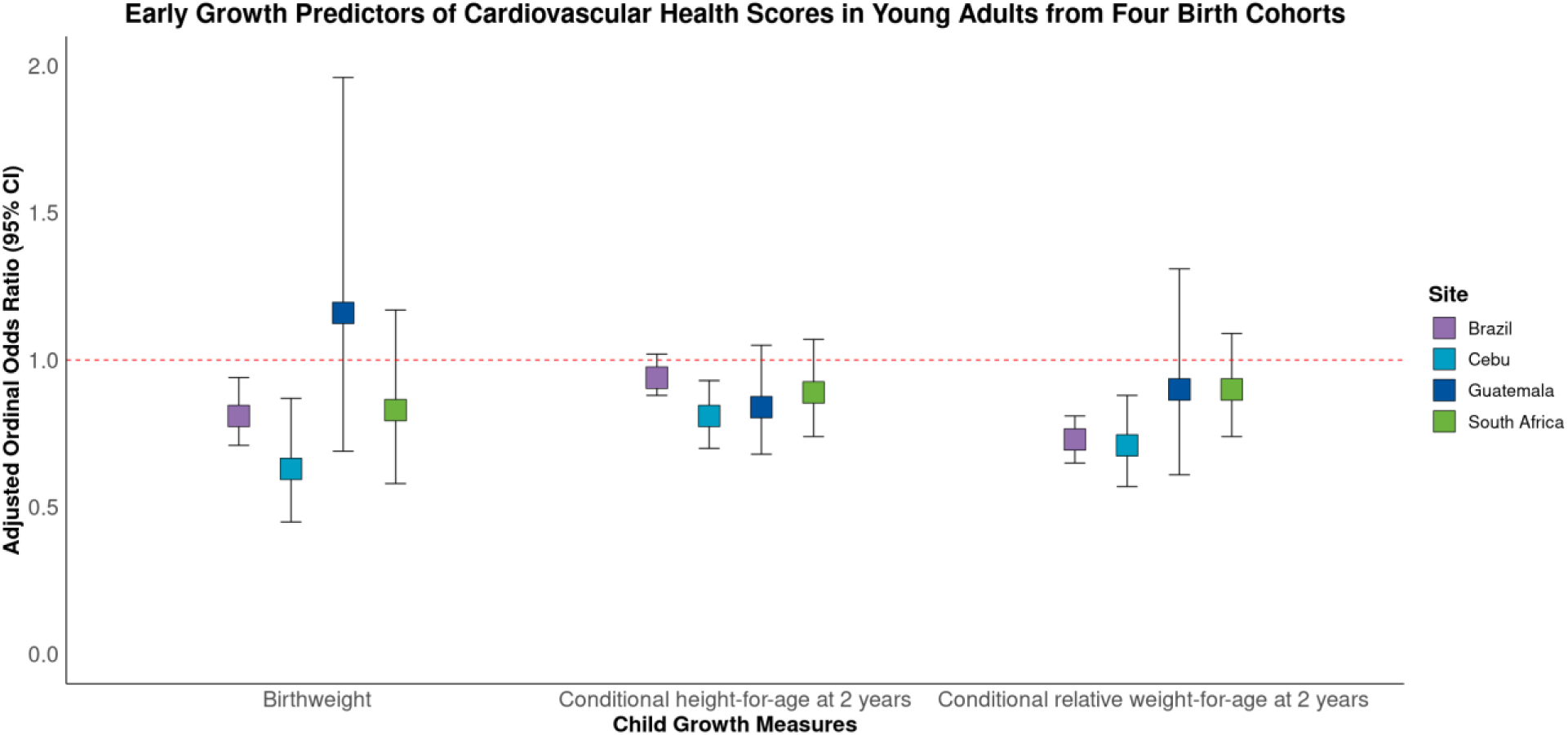
Association of child growth measures with modified Life’s Simple 7 cardiovascular health scores in four COHORTS sites. Ordinal logistic regression model adjusted for maternal/ household factors (maternal age, maternal height, maternal school, child birth order, and household wealth), and potential confounders (child sex, age at adult assessment, attained schooling, and adult IQ).

Exposure and outcome measurements utilized in this study have been described elsewhere (18, 19, 20, 21). Briefly, birthweight was recorded at delivery in hospitals in South Africa and Brazil, while in the Philippines, it was measured either in hospitals or at home by birth attendants using project-provided scales. In Guatemala, a project nurse measured birthweight at home or in a healthcare center. Early life size (length-for-age Z score and weight-for-age Z score) were obtained at 1 year for Brazil and at 2 years for Philippines, South Africa, and Guatemala.

Household wealth was represented by an asset index based on household possessions. Adult weight and height were measured by standard procedures in all sites. Body mass index (BMI) was calculated as weight in kilograms divided by height in meters squared (kg/m^2^). Blood pressure (BP) was measured in Brazil by aneroid sphygmomanometer, in the Philippines by mercury sphygmomanometer, and in Guatemala and South Africa using digital devices UA-767 [A and D Medical] and Omron M6 [Omron], respectively. Fasting blood glucose (FBG) concentration was measured across all cohorts except in Brazil. In Brazil, a random finger-prick capillary whole-blood sample was obtained and adjusted for the time elapsed since the previous meal. In the Philippines cohort, venous blood glucose was assessed using a point-of-care glucometer. To estimate plasma-equivalent glucose levels, a −10% adjustment was applied, aligning with established literature and the methodology used in other COHORTS analyses (12, 22, 23). Adult IQ was measured using the Wechsler Adult Intelligence Scale (3^rd^ edition) in the Brazil cohort, and the Raven’s Standard Progressive Matrices in the Guatemala, South Africa, and the Philippines cohorts. We standardized the distribution within each cohort to a mean of 100 and an SD of 15.

We adjusted for potential confounders including child sex, age at adult assessment to account for age-related variations in the outcomes; number of completed years in school to control for educational attainment’s impact on cognitive and health measures; and adult IQ to account for variations in cognitive ability that could influence the outcomes.

### Measuring cardiovascular health

In 2010, The American Health Association defined CVH based on the Life’s Simple 7 (LS7) metrics (14). The LS7 metrics included diet, physical activity, smoking, BMI, BP, FBG, and total cholesterol. For this study, CVH measures, BMI, FBG, BP, and smoking were available for all four sites. No data on the behavioral factors (diet and physical activity) were available for analysis. We did not use the newer Life’s Essential 8 (LE8) framework because the data available were more consistent with the LS7 metrics criteria for measuring CVH in this context. Therefore, in this analysis, we define CVH as a modified LS7 CVH score. The AHA LS7 definitions of poor, intermediate, and ideal CVH score, along with each component, are presented in Supplementary Table 1 and have been published elsewhere (14).

Using AHA LS7 criteria, components of CVH (i.e., BMI, FBG, BP, and smoking) were allocated a score of 2 for ‘ideal’, score of 1 for ‘intermediate’, and score of 0 for ‘poor’. Age-specific cut-points were used to allocate scores for each LS7 component as needed per the ages at assessment (shown in Supplemental Table 1). The scores were summed to yield the modified LS7 CVH score and were categorized for Brazil, Philippines, South Africa, and Guatemala as ideal (total score 7-8), intermediate (total score 5-6), and poor (total score 0-4). Similar methods have been used previously (15, 24).

### Statistical analysis

We performed bivariate and multivariate ordinal logistic regression analyses to explore the relationship between maternal and child predictors and the odds of having a higher modified total LS7 score or the odds of having a higher score in BMI, FBG, BP and smoking status in the young adult offspring. All models were adjusted for maternal and household factors (maternal age, maternal height, maternal schooling, and household wealth), child size indicators (birth weight, conditional height at 2 years and conditional relative weight-for-age Z scores at 2 years, and birth order), and covariates (age at adult assessment, attained schooling, adult IQ, and child sex). Results are presented as adjusted ordinal odds ratios (AOR) with 95% confidence intervals (CI), and corresponding p-values, such that an AOR above 1.00 is suggestive of a positive association between the exposure and CVH. Descriptive statistics (means and standard deviations for continuous variables; frequencies and proportions for categorical variables) are reported for each site and stratified by sex to examine differences in site and participant characteristics and prevalence of poor, intermediate, and ideal CVH scores. All analyses were performed using SAS software, Version [9.4]

To evaluate whether associations between early-life predictors and later-life cardiovascular health differed by sex, we tested for effect modification by including multiplicative interaction terms between each early-life predictor and sex. Interaction terms were assessed using Wald tests for the cross-product coefficients. Separate models were estimated for total modified CVH score and for each individual LS7 component within each site.

A nominal p-value of <0.05 was considered evidence of potential interaction. To reduce the risk of Type I error from multiple testing across predictors, outcomes, and sites, we applied a Bonferroni correction, yielding an adjusted significance threshold of α = 0.0002.

## RESULTS

### Participant characteristics

Characteristics of participants’ early-life predictors and CVH indicators for each site stratified by sex are described in Table 1. Maternal age was similar across all sites, with a mean of 26 years. Mean maternal height ranged from 148.8 cm to 158.7 cm and maternal schooling ranged from 1 to 10 years. Mean child birthweight was similar between males and females within each site.

**Table 1.** Characteristics of study participants with early-life exposures and cardiovascular health biomarkers from four COHORTS sites.

|  | Brazil |  | Philippines |  | South Africa |  | Guatemala |  |
| --- | --- | --- | --- | --- | --- | --- | --- | --- |
|  | Male (n=1403) | Female (n=1378) | Male (n=580) | Female (n=465) | Male (n=216) | Female (n=243) | Male (n=120) | Female (n=177) |
| **Maternal age (at delivery of index child)** | 26.1 (6.1) | 26.3 (6.4) | 26.5 (6.2) | 26.6 (6.1) | 26.3 (6.4) | 26.1 (6.2) | 27.1 (7.6) | 27.6 (7.3) |
| **Maternal height (cm)** | 156.6 (6.2) | 156.4 (5.9) | 150.4 (4.9) | 150.1 (5.0) | 158.2 (5.7) | 158.7 (6.0) | 148.5 (4.7) | 148.8 (4.9) |
| **Maternal schooling (years)** | 6.5 (4.0) | 6.5 (4.2) | 6.9 (3.2) | 6.7 (3.1) | 9.6 (2.5) | 9.8 (2.4) | 1.3 (1.5) | 1.3 (1.5) |
| **Birthweight (kg)** | 3.3 (0.5) | 3.2 (0.5) | 3 (0.4) | 3.0 (0.4) | 3.2 (0.6) | 3.0 (0.5) | 3.1 (0.5) | 3 (0.4) |
| **Length-for-age Z score at 2 years** | -0.7 (1.2) | -0.6 (1.2) | -2.5 (1.1) | -2.4 (1.1) | -1.4 (1.1) | -1.1 (1.0) | -3.0 (1.2) | -2.9 (1.1) |
| **Weight-for-age Z score at 2 years** | 0.1 (1.1) | 0.1 (1.0) | -1.7 (1.0) | -1.7 (1.0) | -0.6 (1.1) | -0.3 (1.0) | -1.7 (1.0) | -1.6 (1.0) |
| **Birth order** |  |  |  |  |  |  |  |  |
| First | 567 (40.4) | 526 (38.2) | 134 (23.1) | 99 (21.3) | 78 (36.1) | 93 (38.3) | 18 (15.0) | 24 (13.6) |
| Second | 399 (28.4) | 408 (29.6) | 118 (20.3) | 112 (24.1) | 62 (28.7) | 75 (30.9) | 19 (15.8) | 27 (15.3) |
| Third | 229 (16.3) | 220 (16.0) | 110 (19.0) | 83 (17.9) | 40 (18.5) | 33 (13.6) | 12 (10.0) | 25 (14.1) |
| Fourth or later | 208 (14.8) | 224 (16.3) | 218 (37.6) | 171 (36.8) | 36 (16.7) | 42 (17.3) | 71 (59.2) | 101 (57.1) |
| **Family wealth in childhood (Quintiles %)** |  |  |  |  |  |  |  |  |
| First | 267 (19.0) | 245 (17.8) | 99 (17.1) | 87 (18.7) | 26 (12.0) | 36 (14.8) | 21 (17.5) | 41 (23.2) |
| Second | 304 (21.7) | 282 (20.5) | 109 (18.8) | 80 (17.2) | 40 (18.5) | 41 (16.9) | 34 (28.3) | 36 (20.3) |
| Third | 514 (36.6) | 498 (36.1) | 208 (35.9) | 172 (37.0) | 72 (33.3) | 99 (40.7) | 31 (25.8) | 44 (24.9) |
| Fourth | 115 (8.2) | 140 (10.2) | 59 (10.2) | 45 (9.7) | 51 (23.6) | 36 (14.8) | 15 (12.5) | 33 (18.6) |
| Fifth | 203 (14.5) | 213 (15.5) | 105 (18.1) | 81 (17.4) | 27 (12.5) | 31 (12.8) | 19 (15.8) | 23 (13.0) |
| **Adult age (years)** | 22.7 (0.4) | 22.7 (0.4) | 21.5 (0.3) | 21.5 (0.3) | 17.9 (0.4) | 18.0 (0.5) | 30.5 (1.8) | 30.2 (1.7) |
| **Attained schooling (years)** | 11.0 (3.9) | 11.8 (4.2) | 9.6 (3.0) | 10.4 (2.7) | 11.6 (1.5) | 12.0 (1.4) | 5.3 (3.2) | 4.5 (3.0) |
| **Body mass index (kg/m<sup>2</sup>)** |  |  |  |  |  |  |  |  |
| Underweight (<18.5 kg/m<sup>2</sup>) | 12 (0.9) | 31 (2.3) | 48 (8.3) | 77 (16.6) | 18 (8.3) | 5 (2.1) | 1 (0.8) | 0 (0) |
| Normal weight (18.5 - 24.9 kg/m<sup>2</sup>) | 484 (34.5) | 585 (42.5) | 404 (69.7) | 298 (64.1) | 186 (86.1) | 163 (67.1) | 40 (33.3) | 29 (16.4) |
| Overweight (25 - 29.9kg/m<sup>2</sup>) | 571 (40.7) | 421 (30.6) | 100 (17.2) | 70 (15.1) | 9 (4.2) | 45 (18.5) | 61 (50.8) | 68 (38.4) |
| Obese (≥30 kg/m<sup>2</sup>) | 336 (23.9) | 341 (24.8) | 28 (4.8) | 20 (4.3) | 3 (1.4) | 30 (12.4) | 18 (15.0) | 80 (45.2) |
| **Fasting blood glucose (mg/dL)** | 92.2 (30.9) | 86.3 (19.8) | 91.7 (9.7)* | 89.6 (8.4)* | 89.7 (11.1) | 86.2 (10.3) | 114.3 (26.9) | 122.7 (28.2) |
| **Systolic blood pressure (mmHg)** | 127.9 (12.1) | 114.8 (12.0) | 111.9 (11.1) | 100.9 (10.9) | 118.4 (11.0) | 109.1 (11.1) | 122.8 (12.2) | 122.2 (17.4) |
| **Diastolic blood pressure (mmHg)** | 76.9 (9.2) | 74.1 (9.2) | 76.8 (9.5) | 67.4 (9.4) | 74.2 (8.8) | 73.1 (7.8) | 73 (9.0) | 73.9 (11.0) |
| **Smoking status** |  |  |  |  |  |  |  |  |
| Never | 915 (65.2) | 950 (68.9) | 114 (19.7) | 359 (77.2) | 52 (24.1) | 158 (65.0) | 45 (37.5) | 174 (98.3) |
| Former | 126 (9.0) | 117 (8.5) | 193 (33.3) | 77 (16.6) | 37 (17.1) | 35 (14.4) | 44 (36.7) | 2 (1.1) |
| Current | 362 (25.8) | 311 (22.6) | 273 (47.1) | 29 (6.2) | 127 (58.8) | 50 (20.6) | 31 (25.8) | 1 (0.6) |
| **Adult IQ** | 101.0 (15.2) | 99.4 (14.6) | 102.5 (11.1) | 102.0 (11.5) | 100.2 (14.5) | 101.1 (13.7) | 109.1 (15.3) | 99.6 (12.7) |

Note: Data are described as means (SD) or frequency (%). COHORTS, Consortium of Health-Oriented Research in Transitioning Societies; NA, data not available. \* Fasting glucose values for the Philippines site were adjusted using a 10% correction factor.

Children in study sites in the Philippines and Guatemala had lower mean length-for-age and weight-for-age Z scores at 2 years than those from Brazil and South Africa.

At follow-up, the mean age of participants ranged from 18 years in South Africa to 30 years in Guatemala. There was a higher prevalence of overweight and obesity among females than males in Brazil, South Africa, and Guatemala. Mean FBG ranged from ∼88 mg/dL in the Philippines to ∼120 mg/dL in Guatemala and was higher in males compared to females in Brazil (92.2 mg/dL vs 86.3 mg/dL), the Philippines (91.7 mg/dL vs 89.6 mg/dL), and South Africa (89.7 mg/dL vs 86.2 mg/dL). Mean SBP was higher among males compared to females in Brazil (127.9 mmHg vs 114.8 mmHg), the Philippines (111.9 mmHg vs 100.9 mmHg), and South Africa (118.4 mmHg vs 109.1 mmHg). Men reported smoking and higher rates than did women in all sites. By design, mean adult IQ was similar across all sites and between males and females.

### Predictors of cardiovascular health

The prevalence estimates for the modified LS7 total score and its individual components, stratified by sex are presented in Table 2. The proportion of participants with ideal CVH was 11-24% among males and 13-68% among females, while poor CVH was 25-49% among males and 6-43% among females.

**Table 2.** Prevalence estimates for selected Life’s Simple 7 cardiovascular health metrics across four COHORTS sites stratified by sex^a^.

|  | Brazil |  | Philippines |  | South Africa |  | Guatemala |  |
| --- | --- | --- | --- | --- | --- | --- | --- | --- |
| Metric Scores* | Male (n=1403) | Female (n=1378) | Male (n=580) | Female (n=465) | Male (n=216) | Female (n=243) | Male (n=120) | Female (n=177) |
| **Body mass index (kg/m<sup>2</sup>)** |  |  |  |  |  |  |  |  |
| Poor | 336 (24.0) | 341 (24.8) | 28 (4.8) | 20 (4.3) | 3 (1.4) | 30 (12.4) | 18 (15.0) | 80 (45.2) |
| Intermediate | 571 (40.7) | 421 (30.6) | 99 (17.1) | 70 (15.1) | 9 (4.2) | 45 (18.5) | 61 (50.8) | 68 (38.4) |
| Ideal | 496 (35.4) | 616 (44.7) | 453 (78.1) | 375 (80.7) | 204 (94.4) | 168 (69.1) | 41 (34.2) | 29 (16.4) |
| **Fasting blood glucose (mg/dL)\*\*** |  |  |  |  |  |  |  |  |
| Poor | 63 (4.5) | 33 (2.4) | 3 (0.5) | 0 (0.0) | 0 (0.0) | 0 (0.0) | 26 (21.7) | 59 (33.3) |
| Intermediate | 229 (16.3) | 137 (9.9) | 64 (11.0) | 42 (9.0) | 38 (17.6) | 20 (8.2) | 64 (53.3) | 88 (49.7) |
| Ideal | 1111 (79.2) | 1208 (87.7) | 513 (88.5) | 423 (91.0) | 178 (82.4) | 223 (91.8) | 30 (25.0) | 30 (17.0) |
| **Blood pressure (mm/Hg)** |  |  |  |  |  |  |  |  |
| Poor | 120 (8.6) | 33 (2.4) | 23 (4.0) | 3 (0.7) | 62 (28.7) | 42 (17.3) | 5 (4.2) | 19 (10.7) |
| Intermediate | 928 (66.1) | 435 (31.6) | 286 (49.3) | 75 (16.1) | 40 (18.5) | 14 (5.8) | 67 (55.8) | 80 (45.2) |
| Ideal | 355 (25.3) | 910 (66.0) | 271 (46.7) | 387 (83.2) | 114 (52.8) | 187 (77.0) | 48 (40.0) | 78 (44.1) |
| **Smoking status (%)** |  |  |  |  |  |  |  |  |
| Poor | 362 (25.8) | 311 (22.6) | 274 (47.2) | 30 (6.5) | 164 (75.9) | 85 (35.0) | 31 (25.8) | 1 (0.6) |
| Intermediate | 126 (9.0) | 117 (8.5) | 192 (33.1) | 76 (16.3) | 0 (0.0) | 0 (0.0) | 44 (36.7) | 2 (1.1) |
| Ideal | 915 (65.2) | 950 (68.9) | 114 (19.7) | 359 (77.2) | 52 (24.1) | 158 (65.0) | 45 (37.5) | 174 (98.3) |
| **Modified total CVH score\*\*\*** |  |  |  |  |  |  |  |  |
| Poor | 351 (25.0) | 183 (13.3) | 149 (25.7) | 26 (5.6) | 57 (26.4) | 36 (14.8) | 59 (49.2) | 76 (42.9) |
| Intermediate | 711 (50.7) | 581 (42.2) | 330 (56.9) | 122 (26.2) | 127 (58.8) | 87 (35.8) | 48 (40.0) | 77 (43.5) |
| Ideal | 341 (24.3) | 614 (44.6) | 101 (17.4) | 317 (68.2) | 32 (14.8) | 120 (49.4) | 13 (10.8) | 24 (13.6) |

Note: Data are described as n (%). CVH, cardiovascular health; COHORTS, Consortium of Health-Oriented Research in Transitioning Societies. <sup>a</sup> Metrics indicators were limited to those collected at the same time point across all sites; \*Age-specific cut-off points were used to assign scores: poor = 0, intermediate = 1, ideal = 2; \*\* Fasting glucose values for the Philippines site were adjusted using a 10% correction factor; \*\*\* Modified total CVH score categories: ideal = 7-8, intermediate = 5-6, poor = 0-4.

In Brazil, about half of participants had intermediate CVH scores (51% of males and 42% of females). Ideal FBG (79-88%) and smoking scores (65-69%) were common, whereas fewer had ideal BMI (35-45%) or blood pressure (25-66%). In the Philippines, most participants had ideal BMI (78-81%) and FBG (88-91%) scores, and a large majority of females had ideal blood pressure (83%) and smoking status (77%). In South Africa, the majority had ideal BMI (69-94%) and FBG (82-92%) scores, but poor smoking scores were frequent among males (76%). In Guatemala, nearly all females were never-smokers (98%), while fewer males met ideal smoking criteria (38%). Poor FBG and BMI scores were more common among females (33-45%).

Together, these patterns indicate that across LMIC sites, men tend to have lower CVH scores due to higher smoking and blood pressure.

### Associations of child growth with modified Life’s Simple 7 total scores

Table 3 presents the results from the multivariate ordinal logistic regression models for associations between maternal and child factors and the odds of higher CVH scores. Maternal age and height and childhood household wealth were not significantly associated with adult CVH in any site.

**Table 3.** Multivariate ordinal logistic regression results of maternal and child factors associated with higher cardiovascular health in four COHORTS sites.

|  | **Brazil** |  | **Philippines** |  | **South Africa** |  | **Guatemala** |  |
| --- | --- | --- | --- | --- | --- | --- | --- | --- |
|  | **(n=2781)** |  | **(n=1045)** |  | **(n=459)** |  | **(n=297)** |  |
| **Predictors** | **AOR** | **(95% CI)** | **AOR** | **(95% CI)** | **AOR** | **(95% CI)** | **AOR** | **(95% CI)** |
| **Maternal/ Household** |  |  |  |  |  |  |  |  |
| Maternal age (years) | 1.01 | (0.99, 1.02) | 0.98 | (0.95, 1.01) | 0.99 | (0.95, 1.03) | 1.02 | (0.98, 1.06) |
| Maternal height (cm) | 1.00 | (0.99, 1.01) | 1.01 | (0.98, 1.04) | 0.97 | (0.94, 1.00) | 1.00 | (0.95, 1.05) |
| Maternal schooling (years) | 0.99 | (0.97, 1.01) | 0.93** | (0.88, 0.98) | 1.06 | (0.98, 1.15) | 1.00 | (0.85, 1.17) |
| Family wealth (quintile) | 0.93 | (0.87, 0.99) | 1.01 | (0.90, 1.12) | 0.96 | (0.82, 1.12) | 1.15 | (0.97, 1.37) |
| **Child size** |  |  |  |  |  |  |  |  |
| Birthweight (kg) | 0.81** | (0.71, 0.94) | 0.63** | (0.45, 0.87) | 0.83 | (0.58, 1.17) | 1.16 | (0.69, 1.96) |
| Conditional height (24m) | 0.94 | (0.88, 1.02) | 0.81** | (0.70, 0.93) | 0.89 | (0.74, 1.07) | 0.84 | (0.68, 1.05) |
| Conditional relative WAZ (24m) | 0.73** | (0.65, 0.81) | 0.71** | (0.57, 0.88) | 0.90 | (0.74, 1.09) | 0.90 | (0.61, 1.31) |
| **Covariates** |  |  |  |  |  |  |  |  |
| Birth order |  |  |  |  |  |  |  |  |
| First vs second | 0.98 | (0.81, 1.17) | 1.28 | (0.85, 1.93) | 0.85 | (0.53, 1.37) | 1.08 | (0.48, 2.46) |
| First vs third | 0.96 | (0.76, 1.21) | 1.37 | (0.88, 2.15) | 0.88 | (0.47, 1.67) | 1.13 | (0.46, 2.78) |
| First vs fourth or more | 1.02 | (0.78, 1.35) | 1.72 | (1.05, 2.82) | 0.98 | (0.47, 2.04) | 0.74 | (0.30, 1.80) |
| Adult age (years) | 0.84 | (0.69, 1.02) | 0.80 | (0.54, 1.19) | 0.90 | (0.60, 1.33) | 0.96 | (0.85, 1.09) |
| Attained schooling (years) | 1.13** | (1.10, 1.16) | 1.17** | (1.10, 1.24) | 1.07 | (0.94, 1.22) | 1.00 | (0.93, 1.08) |
| Adult IQ (Std. score) | 1.01 | (1.00, 1.01) | 0.99 | (0.98, 1.00) | 1.00 | (0.98, 1.01) | 1.00 | (0.98, 1.02) |
| Adult sex (ref=male) | 0.44** | (0.38, 0.51) | 0.10** | (0.07, 0.13) | 0.26** | (0.18, 0.39) | 0.72 | (0.44, 1.18) |

Note: Outcome is the modified Life's Simple 7 cardiovascular health (CVH) score. COHORTS, Consortium of Health-Oriented Research in Transitioning Societies; AOR, adjusted odds ratio; WAZ, weight-for-age Z score at 2 years. Values shown are from the final model adjusted for maternal/household factors, child size indicators, and other covariates. \*p < 0.05, \*\*p < 0.01.

Birthweight was inversely associated with CVH in Brazil (AOR = 0.81; 95% CI: 0.71, 0.94) and the Philippines (AOR = 0.63; 95% CI: 0.45, 0.87) but not in South Africa or Guatemala.

Similarly, conditional relative weight-for-age 2 years was inversely associated with CVH in both these sites (Brazil AOR = 0.73, 95%CI: 0.65, 0.81; Philippines AOR = 0.71; 95% CI: 0.57, 0.88). Conditional height at age 2 y was inversely associated with CVH in the Philippines (AOR = 0.81; 95% CI: 0.70, 0.93), but not in other sites.

Attained schooling was associated with better CVH in Brazil (AOR = 1.13; 95% CI: 1.10, 1.16) and the Philippines (AOR = 1.17; 95% CI: 1.10, 1.24). No significant association was observed in South Africa or Guatemala.

Sex differences were evident across sites. Female participants had significantly higher odds of better CVH scores compared with males in Brazil (AOR = 0.44; 95% CI: 0.38, 0.51), the Philippines (AOR = 0.10; 95% CI: 0.07, 0.13), and South Africa (AOR = 0.26; 95% CI: 0.18, 0.39), while the estimate was null in Guatemala (AOR = 0.72; 95% CI: 0.44, 1.18).

### Associations of child growth with modified Life’s Simple 7 individual components

Figure 4 presents results of regression analysis of child growth measures and the individual components of the LS7. The strongest associations of child size were with adult BMI scores (Panel A). Higher birthweight was associated with lower odds of healthier BMI in Brazil, the Philippines, and South Africa (all <0.005). Conditional relative weight-for-age at 2 years showed consistent inverse associations with adult BMI scores in all four sites (p<0.01 in Brazil, Cebu, and South Africa; p = 0.003 in Guatemala). Across sites, female participants had lower odds of ideal BMI scores, reflecting the higher rates of overweight or obesity among women, particularly in South Africa (AOR = 9.0; 95% CI: 4.5, 18.1) and Guatemala (AOR = 4.0; 95% CI: 2.4, 6.7).

**Figure 4.**
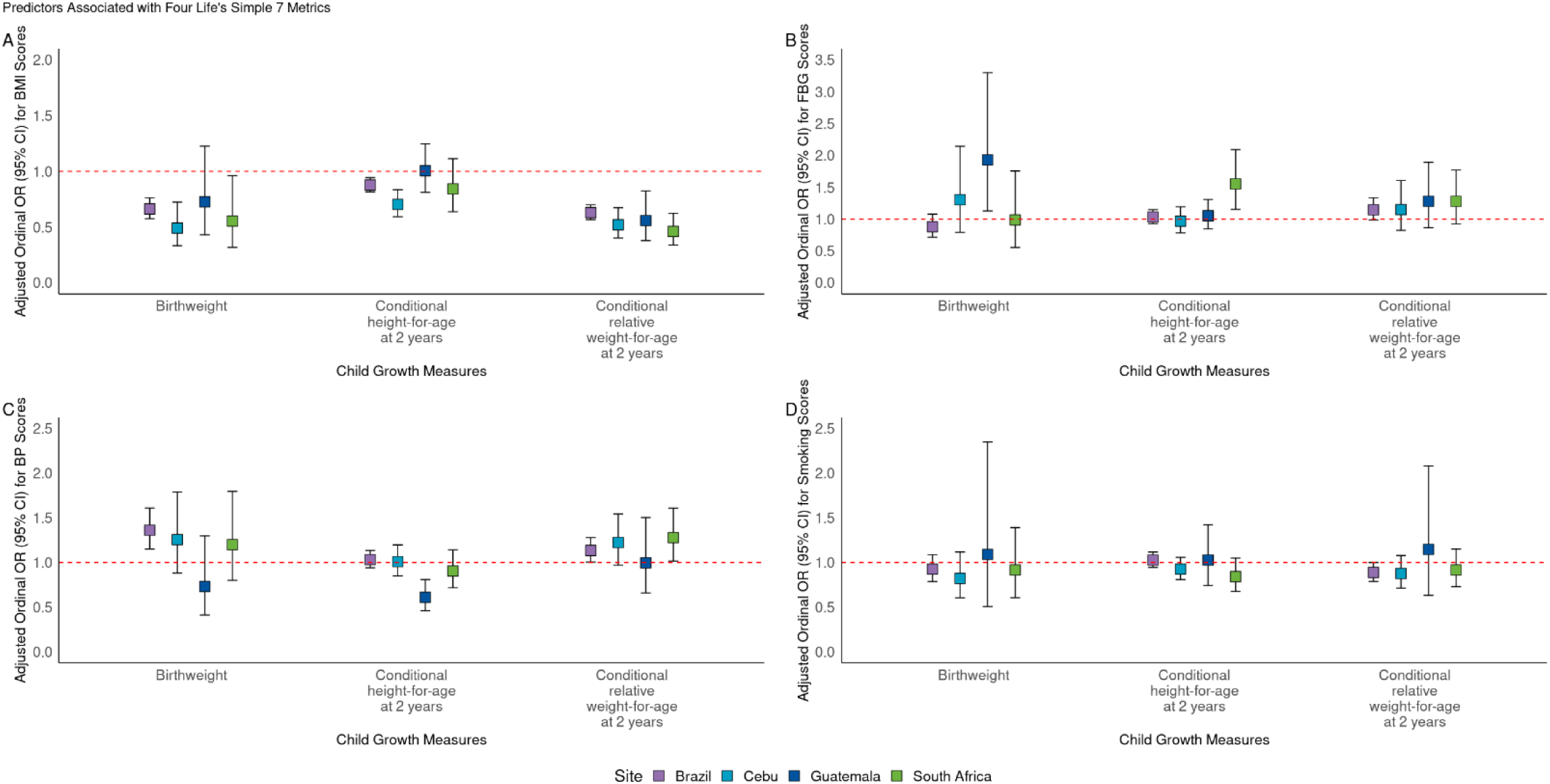
Associations of child growth measures with individual metrics of the modified Life’s Simple 7 scores in four COHORTS sites. A: Odds of having a healthier body mass index (BMI) score. B: Odds of having a healthier fasting blood glucose (FBG) score. C: Odds of having a healthier blood pressure (BP) score. D: Odds of having a healthier smoking status score. Ordinal logistic regression model adjusted for maternal/ household factors (maternal age, maternal height, maternal school, child birth order, and household wealth), and potential confounders (child sex, age at adult assessment, attained schooling, and adult IQ. FBG model controlled for adult BMI, and BP model controlled for adult height and BMI

Birthweight was positively associated with healthier FBG scores in Guatemala only (AOR = 1.93; 95 % CI: 1.13, 3.30) after controlling for adult BMI (Panel B). Across most sites, men had higher odds of poor FBG scores than women, particularly in Brazil (AOR = 0.55; 95% CI: 0.45, 0.69), South Africa (AOR = 0.41; 95% CI: 0.21, 0.81), and Guatemala (AOR = 0.91; 95% CI: 0.86, 0.95).

Child size indicators were positively associated with healthier BP scores in young adulthood across sites after controlling for adult height and BMI (Panel C). Birthweight was positively associated with healthier adult BP scores, reaching statistical significance in one of three sites (Brazil AOR = 1.36; 95% CI: 1.15, 1.61). Conditional relative weight at age 2 years was positively associated with healthier BP scores, reaching statistical significance in two (Brazil: AOR = 1.13; 95% CI: 1.01, 1.28 and South Africa AOR = 1.28; 95% CI: 1.01, 1.61) of four sites. Across all sites, females participants had higher odds of ideal BP: Brazil (AOR = 0.22; 95% CI: 0.16, 0.29), Philippines (AOR = 0.20; 95% CI: 0.12, 0.33), South Africa ( AOR = 0.26; 95% CI: 0.13, 0.52), and Guatemala (AOR = 0.28; 95% CI: 0.11, 0.76).

Child growth measures and adult smoking was unrelated across all sites (Panel D). In contrast, higher educational attainment was strongly associated with better (non-smoking) scores in Brazil, the Philippines, and South Africa (all p<0.005). Men were substantially more likely to report current smoking status than women in every site, consistent with the large sex gap in smoking prevalence observed in Table 2.

In secondary analyses, we tested sex interaction to assess whether the associations between early-life exposures and adult CVH differed for men and women. Although a few interaction terms reached significance, none remained significant after Bonferroni correction across all exposures, outcomes, and sites (α = 0.0002). Thus, there was no consistent evidence that the effects of early-life predictor on adult CVH differed by sex.

## DISCUSSION

This study provides novel insights into the long-term associations of early-life exposures, including maternal and household factors and child growth, with cardiovascular health (CVH) in young adulthood in four low- and middle- income (LMIC) countries. Using harmonized data from the COHORTS collaboration, we demonstrated that greater early-life weight and faster weight gain in the first two years of life were associated with lower young-adult CVH scores, particularly in Brazil and the Philippines cohorts. Attained schooling was positively associated with better CVH scores in both sites. Across most sites, males had lower odds of ideal CVH, driven largely by lower blood pressure and smoking scores. These findings underscore the importance of early growth, education, and sex-specific vulnerabilities in shaping cardiovascular trajectories in LMICs.

The strong links between early growth and CVH align with extensive evidence that the first 1,000 days of life are critical for long-term cardiometabolic health (5, 6, 7, 13, 24). The present analysis extends this body of work by examining multiple birth cohorts from different regions and examining a composite CVH metric and specific CVH components, providing new insights into the long-term health biomarkers across different populations. Previous research has demonstrated that early life adversities, such as poor growth (low birthweight, slow linear growth, rapid weight gain), are associated with specific cardiometabolic biomarkers, such as elevated body mass index (BMI), blood pressure (BP), and impaired glucose metabolism (12). Our study builds on this evidence by integrating these individual risk factors into a composite measure of CVH using the American Heart Association’s (AHA) Life’s Simple 7 (LS7) metrics. This life-course perspective highlights how early-life exposures translate into measurable cardiovascular differences in young adulthood and reinforces the need for interventions during early childhood.

The positive association between educational attainment and CVH observed in Brazil and the Philippines highlights the potential role of education in promoting long-term health. Education has long been recognized as a determinant of health, influencing health behaviors, access to care, and economic opportunities (25, 26). In Guatemala and South Africa, the association between education and CVH score did not reach statistical significance, likely attributable to sample size limitations. Additionally, relatively uniform educational attainment or limitations in access to education could limit our ability to assess the relationship between education and CVH. A summary of studies suggest that investments in educational quality and duration can serve as a primordial prevention strategy for cardiovascular diseases, thereby improving population health outcomes (27). This aligns with the notion that education influences health literacy, which in turn effects health behaviors and outcomes. Individuals with higher educational attainment are more likely to engage in health-promoting behaviors, such as healthy eating and regular exercise, which are crucial for maintaining CVH (27). One study found the low prevalence of high CVH in Peru underscored the need for targeted interventions that consider educational background as a key factor in health promotion strategies (28). Our findings highlight the importance of context-specific strategies to enhance educational opportunities, particularly in settings where access remains limited.

Higher birthweight and greater conditional relative weight at 2 years were associated with lower odds of having an ideal BMI in adulthood. This aligns with studies showing that rapid early weight gain, especially when disproportionate to linear growth, can increase the risk of overweight and obesity later in life (7, 29). In contrast, higher birthweight was linked to healthier blood pressure scores in adulthood, reflecting lower odds of elevated BP. Previous research has also found that higher birthweight was associated with reduced odds of elevated BP in adulthood (12). This supports the developmental origins of health and disease (DOHaD) framework, which posits that fetal undernutrition and restricted intrauterine growth may program long-term vascular and renal changes that increase susceptibility to hypertension (30). Adequate fetal growth, reflected in higher birthweight within the normal range, is thought to promote healthier vascular development and more favorable regulation that contribute to more optimal blood pressure in later life (30). Together, these findings highlight a trade-off in early growth patterns: higher birthweight and early weight gain may improve blood pressure outcomes yet simultaneously increase the risk of adult overweight. This duel influence emphasized the need for interventions that promote healthy early growth to support development but without encouraging excessive adiposity (31).

Our study is among the first to examine the association of early-life factors on a composite CVH score in LMICs, integrating BMI, BP, fasting glucose and smoking to assess CVH later in life.

This novel approach offers a life course perspective that underscores critical windows for intervention and the importance of early prevention strategies. Also, although the CVH score was constructed using standardized criteria across sites, associations between early-life exposures and later-life CVH were weaker in the South African cohort compared to other COHORTS sites and prior studies. While we applied adult blood pressure thresholds for participants aged 17 and older, consistent with American Academy of Pediatrics and AHA LS7 guidelines, differences in the overall CVH score construction, particularly the limited availability of behavioral components such as diet and physical activity, may have influenced how cardiovascular health was captured (32). Moreover, previous studies of COHORTS data focused on individual risk factors, such as body composition, diabetes, and BP (9, 12, 29, 33). The innovation in our study lies in examining how these measures collectively reflect CVH, offering a broader understanding of the long-term impacts of early growth patterns. The general findings provide a foundation for further research into the mechanisms linking early-life growth to overall CVH.

Sex differences were consistent across sites, with males demonstrating poor overall CVH, possibly due to observed higher smoking prevalence and elevated BP. However, formal tests of interactions showed no consistent modification of early-life associations by sex after correction for multiple testing. This suggests that while men and women differ in overall CVH levels, the pathways linking early growth to later cardiovascular outcomes are similar in both sexes.

This study has several strengths. First, the general consistency of our findings across multiple LMIC sites enhances the transportability of the results to other similar settings. By leveraging prospective birth cohort data, our study provides robust evidence of the long-term effects of early-life exposures on CVH. The inclusion of distinct samples from three continents allows us to account for regional variations in socioeconomic conditions and health systems. It is important to acknowledge that while the COHORTS datasets offer rich longitudinal insights, they are not population-based, and the findings may not be fully generalized to national populations. Nonetheless, they provide critical insights into the long-term impacts of early-life exposures in LMICs.

Limitations of our study should be noted. First, the age of the data, which may not fully reflect contemporary developments in these countries. Over the past few decades, socioeconomic conditions, healthcare access, and educational opportunities have changed, potentially altering the associations observed in our study. Nevertheless, the findings remain relevant as they highlight the lasting effect of early-life factors on CVH, a relationship unlikely to change significantly over time. Additionally, the absence of diet, physical activity, and sleep data prevent us from implementing the full AHA LS7 or Life’s Essential 8 (LE8) metrics potentially limiting construct validity. Despite this limitation, our modified total score provide a meaningful measure of CVH, align with prior research in similar contexts, and may in fact be underestimating true CVH (15). Therefore, future research using individual datasets could leverage additional variables from newer waves of data collection to examine cardiovascular health within the LS7 or the more recent LE8 framework.

In conclusion, our findings emphasize that early-life factors, particularly during the first 1,000 days, and child growth patterns significantly predict CVH in young adulthood. The associations observed emphasize the importance of early interventions in potentially influencing lifelong cardiovascular biomarkers and outcomes. Investments in maternal and child health programs, including improved access to nutrition, education, and healthcare, are critical for addressing health disparities in LMICs (7). Strategies that promote health growth and educational attainment during early life have the potential to reduce the burden of cardiovascular disease in later adulthood (8, 25). By adopting a life-course approach starting in early childhood, public health initiatives can target the roots of cardiovascular disease and contribute to improving global cardiovascular health.

## Data Availability

The data that support the findings of this study are available from the corresponding author upon reasonable request.

## ACKNOWLEDGMENTS

We would like to acknowledge the dedicated members of the COHORTS project team and data collectors, and the parents and children who participated for their invaluable contributions to the project.

## Supplementary Material

**Table S1.** Selected American Heart Association Life’s Simple 7 metrics definitions of poor, intermediate, and ideal cardiovascular health.

| **Goal/ Metric** | **Poor**<br>Definition | **Intermediate**<br>Definition | **Ideal**<br>Definition |
| --- | --- | --- | --- |
| **Current smoking** |  |  |  |
| Adults >20 years | Yes | Former ≤12 months | Never or quit >12 months |
| Children 12–19 years | Tried prior 30 days | - | Never tried; never smoked whole cigarette |
| **Body mass index** |  |  |  |
| Adults >20 years | ≥30 kg/m<sup>2</sup> | 25–29.9 kg/m<sup>2</sup> | <25 kg/m<sup>2</sup> |
| Children 12–19 years | >95th Percentile | 85th–95th Percentile | <85th Percentile |
| **Total cholesterol** |  |  |  |
| Adults >20 years | ≥240 mg/dL | 200–239 mg/dL or treated to goal | <200 mg/dL |
| Children 12–19 years | ≥200 mg/dL | 170–199 mg/dL | <170 mg/dL |
| **Blood pressure** |  |  |  |
| Adults >20 years | SBP ≥140 or DBP ≥90 mm Hg | SBP 120–139 or DBP 80–89 mm Hg or treated to goal | <120/<80 mm Hg |
| Children 12–19 years | >95th Percentile | 90th–95th Percentile or SBP ≥120 or DBP ≥80 mm Hg | <90th Percentile |
| **Fasting plasma glucose** |  |  |  |
| Adults >20 years | ≥126 mg/dL | 100–125 mg/dL or treated to goal | <100 mg/dL |
| Children 12–19 years | ≥126 mg/dL | 100–125 mg/dL | <100 mg/dL |

Notes: Other metrics included in Life's Simple 7 that are not included in the table are physical activity and healthy diet score. SBP, systolic blood pressure; DBP, diastolic blood pressure

